# Rabies mortality and morbidity associated with animal bites in Africa: A case for Integrated Rabies Diseases Surveillance, Prevention and Control - A Scoping review

**DOI:** 10.1101/2020.07.29.20164483

**Authors:** Peter Nyasulu, Jacqueline Weyer, Rea Tschopp, Adane Mihret, Abraham Aseffa, Samuel Victor Nuvor, Tolulope Balogun, Jacques Lukenze Tamuzi, Luke Nyakarahuka, Gideon Kofi Helegbe, Nyanda Elias Ntinginya, Melaku Tefera Gebreyesus, Seydou Doumbia, Lucille Blumberg, Reinhard Busse, Christian Drosten

**Affiliations:** Division of Epidemiology & Biostatistics, Department of Global Health, Faculty of Medicine & Health Sciences, Stellenbosch University, Cape Town, South Africa; Centre for Emerging Zoonosis and Parasitic Diseases, NICD, NHLS, Johannesburg, South Africa; Swiss Tropical and Public Health Institute, Socinstr. 57, 4002 Basel, Switzerland; University of Basel, Basel, Switzerland; The Armauer Hansen Research Institute (AHRI), PO Box 1005, Addis Ababa, Ethiopia; Department of Microbiology and Immunology, School of Medical Sciences, University of Cape Coast, Cape Coast, Ghana; Department of Biosecurity, Ecosystems and Veterinary Public Health, Makerere University, Kampala, Uganda; Department Biochemistry and Molecular Medicine, School of Medicine and Health Sciences, University for Development Studies, Tamale, Ghana; Mbeya Medical Research Centre, National Institute of Medical Research, Tanzania; Department of Veterinary Medicine, Lilongwe University of Agriculture and Natural Resources, Lilongwe, Malawi; University Clinical Research Center, Faculty of Medicine, University of Bamako, Mali; Institute of Virology, Charité Global Health, Charité - Universitätsmedizin Berlin, Campus Charité Mitte, Germany

**Keywords:** rabies, mortality, morbidity, surveillance, zoonosis, neglected tropical disease, Africa

## Abstract

**Background:** Rabies a neglected tropical disease, mostly affecting poor and vulnerable populations living in remote rural areas in developing countries. The disease continues to pose a significant public health a threat with an estimated 59,000 dog-transmitted human deaths, of which an estimated 21,476 human deaths occur in Africa each year. The global strategy has been set by the quartite World Health Organization (WHO), the World Organization for Animal Health (OIE), the Food and Agriculture Organization of the United Nations (FAO) and the Global Alliance for Rabies Control (GARC), aiming for “zero human deaths associated with dog transmitted rabies by 2030”. African countries, however, face several challenges and there are still gaps in controlling rabies. The aim of this study review is to determine rabies prevalence, mortality and associated risk factors in both human population and animal population and to evaluate the presence or absence of integrated one health surveillance response in African nations.

**Methods and analysis:** We will conduct an electronic literature searches on PubMed, CINAHL, (Cumulative Index of Nursing and Allied Health Literature), Scopus, and Web of Science and other relevant databases. Reference lists from identified published articles or reviews and conference abstracts will also be searched for relevant articles. Published and unpublished literatures (grey) will be included in the search. The findings will be presented graphically in terms of mortality, morbidity, interventions for rabies control in Africa, research gaps identified, available research evidence, rabies surveillance, prevention and control and adverse events.

**Conclusion:** This review will contribute to the coordination of interventions for surveillance, prevention, and control with African countries as country-based gaps and challenges and opportunities will be highlighted. In addition, the scaling up of post-exposure prophylaxis (PEP rabies will be evaluated in African countries and the projection to achieve the target of “zero deaths of human rabies by 2030” in Africa.

**Ethics and dissemination:** Since this is a scoping review, there is no institutional requirement to obtain ethical clearance from the Health Research Ethics Committee of the Faculty of Medicine and Health Sciences, Stellenbosch University. Data used will be from open source and publicly available accessed on different databases as described in the methods. Ethical conduct of scientific investigator regarding publication of the work arising from this protocol will be adhered to by the research team. The findings will be disseminated in clinical seminars, scientific forums and conferences targeting infectiology for human and veterinary clinician, public health scientists, virologists, immunologists, vaccinologists and policy makers among others.

*http://creativecommons.org/licenses/by-nc/4.0/*

*This is an open access article distributed in accordance with the Creative Commons Attribution Non Commercial (CC BY-NC 4.0) license, which permits others to distribute, remix, adapt, build upon this work non-commercially, and license their derivative works on different terms, provided the original work is properly cited, appropriate credit is given, any changes made indicated, and the use is non-commercial*

**Strengths and limitations of the study:**

- This is the first scoping review to synthesize new and old evidences on rabies and weight those evidence to achieve global target of “zero human rabies deaths by 2030” in Africa.
- The review will include published and non-published literature (i.e. peer reviewed and non-peer reviewed) in different African countries. So called grey literature is key in a review of this topic as surveillance data and other related findings are often not reported in peer-reviewed scientific journals.
- Mapping outcomes are designed as evidence-based practices that may play key role in African countries rabies public health policy.
- As with most scope reviews, this scope review does not assess the quality of evidence and the risk of bias in various studies. In addition, the review will include a large number of studies (both published and unpublished), which will require considerable time to complete the study.

## Background /Introduction

Rabies is a zoonotic disease caused by a *Lyssavirus*, belonging to the family Rhabdoviridae. Clinically the infection manifests itself as a fulminating encephalomyelitis. Rabies is one of the neglected tropical diseases mainly affecting poor and vulnerable communities living in remote rural areas in mostly developing countries [1]. The disease remains a threat to public health worldwide with approximately 15.69 million people potentially exposed to the virus every year, requiring post-exposure prophylaxis (PEP) and an estimated 59,000 deaths [2]. Rabies occurs on all continents, with more than 95 per cent of human deaths occurring in Asia and Africa [2]. An estimated 21,476 human deaths in Africa from dog-mediated rabies occur every year [2]. Approximately 80 percent of human cases occur in rural areas [1] and children between the ages of 5-14 are frequent victims [2]. Today, rabies-free regions or countries in mainland Africa are not known [3]. Athough there are successful human and immunoglobulin vaccines for rabies, they are not readily available or open to those in needs [2]. It is estimated that Africa spends the least on PEP, and has the highest human mortality cost [2]. WHO, the United Nations Food and Agriculture Organization (FAO), the World Organization for Animal Health (OIE), and the Global Alliance for Rabies Control have set a global target of “zero human rabies deaths by 2030” [1]. With improved dog rabies immunization programs to reduce rabies in domestic dogs and enhanced access to PEP a significant number of human lives could be saved.

Reviewing data from different African countries, the literature showed that domestic dogs’ rabies is endemic, and cases in different communities have been neglected [4]. From 2010 to 2014, 22 cases of human rabies and dog bites were reported at the Korle Bu Teaching Hospital and 233 dog bites at Ridge Hospital in Accra, the capital city of Ghana [5]. In Ghana’s eastern region, 4821 dog bites were reported over a three-year period, with 53.3 percent of cases of rabies recorded in seven of the 26 municipalities and districts [6]. In Uganda, through a retrospective data analysis at the Ministry of Health, a total of 117,085 cases of rabies and 371 deaths were registered over a nine-year period (2001-2009) [7]. In addition, from 2001 to 2015, another study reported 208,720 cases of animal bites in the Uganda Ministry of Health database [8]. A recent study by Uganda’s Ministry of Agriculture recorded 8,240 animal bites and 156 deaths from 2013-2017 suspected of being caused by the rabies virus (Mwonje et al, unpublished data). In Uganda, victims of dog bites account for 96 per cent of cases of animal bite with an estimated caseload of 6,601.5 cases per year and an incidence of 39.6/100,000 people [9]. The mortality rate from rabies is estimated at 0.30/100,000 inhabitants [9]. The demand for PEP rabies to victims of animal bite is generally high and some of the reported cases are not true rabies cases. However, it remains unclear whether dog bites are effectively managed with adequate anti-rabies PEP vaccine and whether individuals exposed to rabies virus by animal bite may die at home without knowing the cause of death as autopsy is not routinely done to determine the cause of death. A study in the Democratic Republic of Congo has shown that rabies in Kinshasa is endemic where dogs and cats played a major role as carriers and transmitters of the rabies virus [10]. There were a total of 5,053 attacks in the veterinary clinics that inflicted either bite or scratch on humans and other animals [10]. Of these, 2.5% were found to be rabid on the basis of clinical observation in 128 cases [10]. Nevertheless, out of 44 brain samples received and examined in laboratories, 29 were found positive for rabies, accounting for 65.9 percent as a positive proportion of the specimens submitted by the laboratory [10]. Data showed the burden of rabies remains scanty in Tanzania due to lack of a formal monitoring mechanism [11]. Dog-bite fatalities estimated annual mortality of 1,499 human rabies per year, equivalent to an annual incidence of 4.9 deaths/100,000 when active bite monitoring data were used, and 193 deaths per year, corresponding to an annual incidence of 0.62 (0.1-1.32) deaths/100,000 when using national bite statistics[11]. Another research conducted in four districts in Tanzania reported an annual rabies incidence rate of up to 35 per 100,000 people and death by verbal autopsy of up to 3.4 per 100,000 persons [12]. An estimated 500 people die of rabies in Malawi annually [13]. However, as most cases remain undiagnosed, hence untreated and unreported, there is potential for underestimating rabies [13]. This paradigm is motivated by the “absence of a national rabies control plan, insufficient laboratory facilities, and lack of supervisory system [13]. In addition, the burden of human rabies from dog bites is on the rise due to poor intersectoral cooperation between human and animal health in Malawi [13]. That was a cause for low annual coverage of rabies vaccination. Just 0.5 per cent was vaccinated with an estimated 400,000 dogs in the country [13]. A retrospective study examining laboratory-based data over a 25-year period, established in South Africa about 353 confirmed cases of human rabies [14]. Of these, 79 percent (n=279) were registered from the province of KwaZulu Natal, and 8 percent (n=29) from the Eastern Cape [14]. The rest, from the other provinces, were published. Overtime occurrence of rabies in KwaZulu Natal has dropped significantly, with less than 10 cases reported in 1995[14]. Other provinces at the same time showed a steady increase in cases of rabies. Over 70% of reported cases of rabies in the childhood population occurred in all of this [14].

Reviewing the above literature, it is clear that data describing the magnitude of rabies in Africa are lacking. This drives the need for a scoping review to establish the landscape of rabies morbidity and mortality in the African Continent and create that as a foundation for establishment of a surveillance program in the continent.

## History of Rabies in Africa

The history of rabies in Africa is not well recorded, but it is well accepted that the disease must have been present for hundreds of years in northern Africa, especially as an urban dog disease and also associated with cycles in the Middle East [3]. In many sub-Saharan African countries, rabies only became epizootic during the nineteenth and twentieth centuries; in this region, the disease became well established among dogs and involved wildlife species over large areas[3-15]. Moreover, Africa harbors several viruses linked to rabies. Historically, isolation and epidemiological analyzes of these viruses have largely been linked to specific surveillance studies or diagnostic competencies [3-16].

## Rabies in Domestic and Wild Animals

Rabies virus spreads in terrestrial hosts in Africa and was not related to infections in bats. Although all mammals are vulnerable to rabies virus infection, some are able to maintain certain variants of the virus adapted to their species [17]. For example, in South Africa, both domestic and wild carnivore species are often diagnosed with infection with the rabies virus [17]. While most of the confirmed cases of rabies in wild carnivorous species are recorded from yellow mongoose (*Cynictis penicillata*), the remaining cases originated from other wild carnivores, as well as from the critically endangered wild dog (*Lycaon pictus*) [18]. In 2014 and 2015, infection with lyssavirus was found in two wild dogs and a spotted hyaena (*Crocuta crocuta*) in the Madikwe Game Reserve, North West Province of South Africa, using a direct fluorescent antibody and immunohistochemistry test [18].

## Rationale for the study

Despite the apparent burden of rabies in Africa, insufficient investment has been made on researching the problem. This is evident in the great lack of published findings on this issue. Many thousands of deaths from rabies are observed in humans worldwide annually, but most of the cases recorded are highly underestimated [19]. Surveillance, prevention and control of rabies diseases in many African countries are very poor or inexistent. Rabies is considered under-reported in many countries, partially due to a lack of surveillance and laboratory materials [20]. Cultural or social taboos further complicate this [21]. In addition, the lack of reliable data on disease occurrence often limits policy makers and public health practitioners’ prioritization of rabies research [22]. It is therefore important to undergo this scoping review to have a clearer picture of the research efforts done so far on rabies in Africa and what can be done to increase awareness and involvement of research and public health practitioners. The review will help identifying any gaps and challenges (depending on the country) in surveillance and control that could be targeted in future programs/research. The one health surveillance approach with an integration of animal and human health practitioners is missing in many African countries. This review will help to identify existing one health disease surveillance in Africa.

## Scoping Review Question

Our review research question is elaborated below using the ‘PICO’.

### Population

Humans infected with rabies **in** African countries.

### Interventions

Integrated Rabies Disease Surveillance, prevention and control.

### Comparisons

Little or no integrated rabies disease surveillance, prevention and control.

### Outcomes

Reduced human morbidity and mortality of rabies associated with animal bites.

### Research question

Would an integrated rabies disease surveillance, prevention and control be effective in reducing the morbidity and mortality of rabies associated with animal bites compared to little or no integrated rabies disease surveillance, prevention and control?

### Aim

The aim of this scoping review is to determine the prevalence, mortality and related risk factors associated with rabies in the human population as well as animal population and assess the presence or absence of one health surveillance-responses in African countries.

### Objectives

To explore the above aim, we will consider the following objectives:

1. To assess the extent of available research on the morbidity and mortality of rabies due to animal bites conducted in Africa.
2. To identify research gaps in the literature on the impact of rabies in Africa so as to effectively plan effective public health intervention.
3. To ascertain the current level of rabies disease surveillance, prevention and control that exist in African countries.
4. To assess the adverse events and complications associated with rabies vaccination in African countries.
5. To assess the different types of vaccines used and the effectiveness of locally produced and imported vaccines in treating rabies in different parts of Africa.
6. To demonstrate that rabies research has been inadequate and should be of public health significance which needs further research in Africa.
7. To assess rabies morbidity and mortality associated with dog and livestock bites in humans and animal population.
8. To summarize and disseminate research findings on rabies in Africa in order to sensitize researchers and policy makers to rise to the challenge of combating rabies.

## Methods

### Types of participants

Studies which include men and women, young and old infected or affected with rabies in Africa will be considered.

### Outcome

The primary outcome measure will be mortality in humans due to rabies as a result of animal bites. Secondary outcomes will include adverse events and morbidity, different methods of rabies disease surveillance, prevention and control measures in different African countries.

### Search strategy

The researchers will develop a comprehensive search strategy with relevant search terms. The researcher will search databases such as PubMed, CINAHL, (Cumulative Index of Nursing and Allied Health Literature), Scopus, and Web of Science and other relevant databases. Reference lists from identified published articles or reviews and conference abstracts will also be searched for relevant articles. Published and unpublished literatures (grey) will be included in the search.

### Reporting of the review findings

The scoping-systematic review protocol will be developed using the methodology of the Joanna Briggs Institute (JBI) [23]. The protocol will be registered in the registry of systematic reviews at Prospero. The scoping review findings will be reported using the Systematic Review and Meta-Analysis Preferred Reporting Items (PRISMA-2009) [24].

### Inclusion criteria

- All studies conducted in the last 10 years or including sites in Africa reporting rabies infections in humans and animals will be included.
- All studies reporting morbidity and mortality of rabies in male and female humans will be included in the review.
- All studies reporting Rabies Diseases Surveillance, Prevention and Control in Africa will be included.
- Cross sectional, case control and cohort studies and possible quasi experimental designs and randomized controlled trials carried out in African countries reporting rabies treatment and control.

### Exclusion criteria

- All studies conducted using data from out outside of Africa will be excluded.
- Non-English articles will be excluded from the review.

### Selection of studies

The search results will be independently screened by two researchers using the abstracts. The screened abstracts will be used to retrieve full articles based on the inclusion and exclusion criteria. The preferred reporting items for systematic reviews and meta-analyses (PRISMA) checklist will inform the selection of studies [24].

### Extracting and charting the data

Data will be extracted from selected studies by two independent researchers using a standardized data extraction form. The extracted data will include information such as author, journal, country of study, study design and year of publication. The researchers will extract primary study data reported for each of the included studies on study design, population, and rabies morbidity and mortality as well as other inclusion criteria parameters. In case of disagreements, a third researcher will be contacted for confirmation.

**Figure 1:**
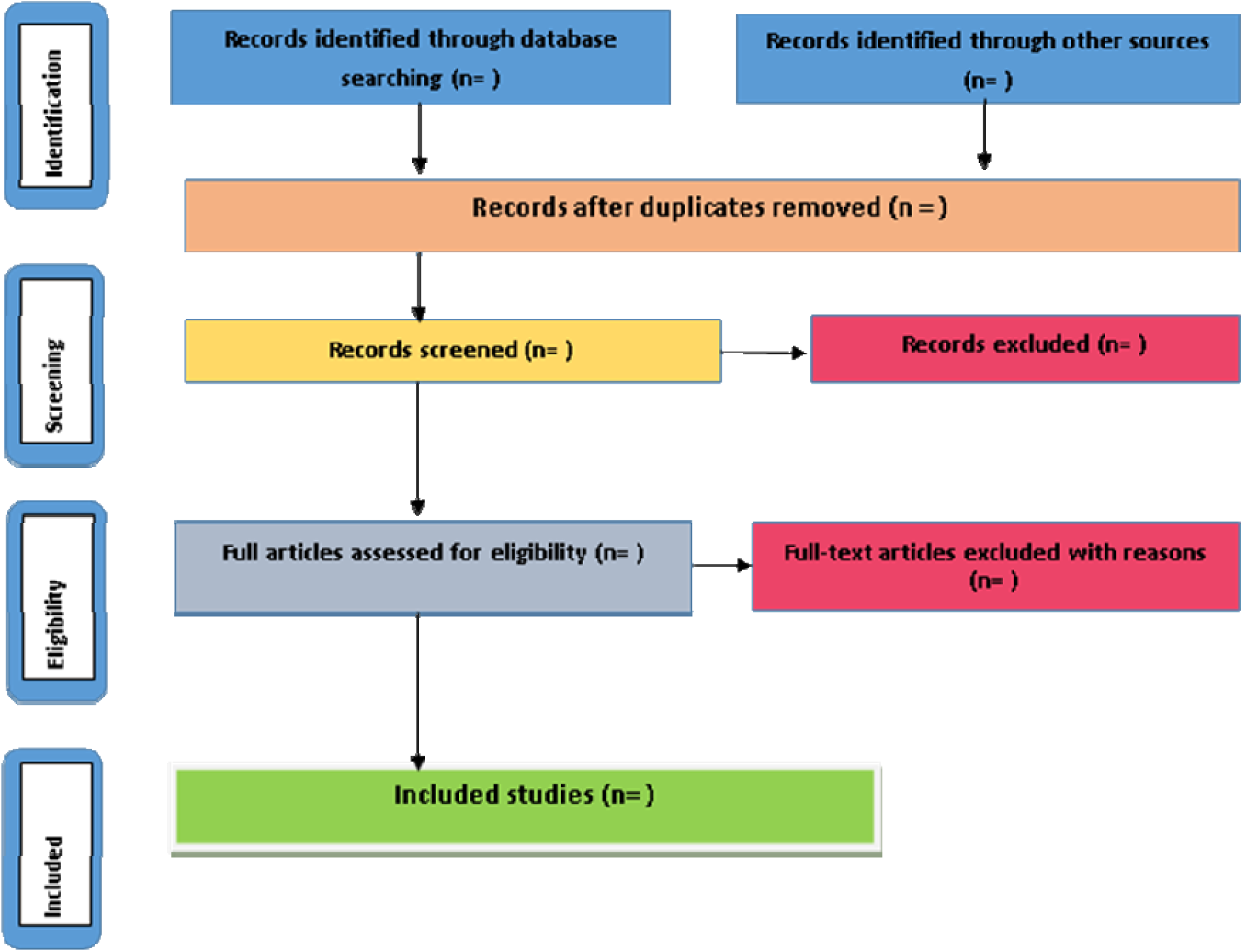
Preferred Reporting Items for Systematic Reviews and Meta-Analysis (PRISMA) flow diagram for the scoping review process [25]

## Mapping of the review findings

The outcome or the findings from the review will be presented graphically.

**Figure 2:**
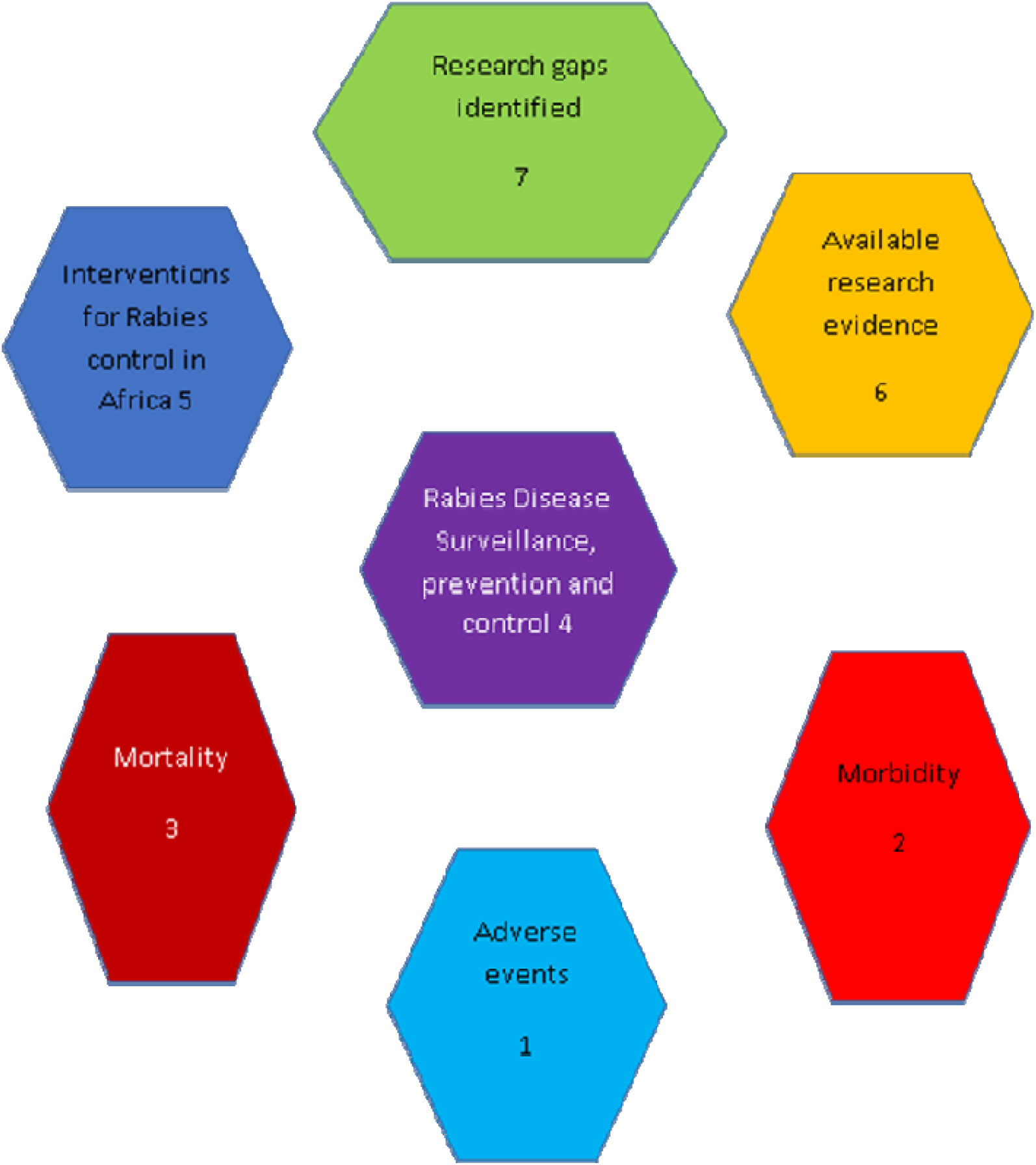
Map of outcome findings from the review the study.

## Review implications

This review will play a substantial role in coordinating rabies related interventions (surveillance, prevention and control) in African countries as gaps and challenges country-based will be highlighted. Additionally, scaling up rabies PEP will be evaluated in African countries and the projection to achieve the target of “zero human rabies deaths by 2030” in Africa.

## Data Availability

No data available yet as this is a proposed study

## Acknowledgments

not applicable

## Ethical considerations

Not applicable

## References

1. World Health Organization. Rabies. https://www.who.int/news-room/fact-sheets/detail/rabies. 21 April 2020. Accessed 10 June 2020

2. World Health Organization. Rabies. https://www.afro.who.int/health-topics/rabies. 2017.

3. Nel Louis H. Discrepancies in data reporting for rabies, Africa. Emerging infectious diseases 2013;19(4):529–33. Accessed 10 June 2020.

4. Apanga PA, Awoonor-Williams JK, Acheampong M, Adam MA. A presumptive case of human rabies: a rare survived case in rural Ghana. Frontiers in public health 2016;4:256.

5. Elieza S. Trends in Dog Bites and Human Rabies in Greater Accra Region, Ghana. http://ugspace.ug.edu.gh/handle/123456789/8470 2016.

6. Adomako BY, Baiden F, Sackey S, Ameme DK, Wurapa F, Nyarko KM, et al. Dog Bites and Rabies in the Eastern Region of Ghana in 2013–2015: A Call for a One-Health Approach. Journal of tropical medicine 2018; 2018:7–12.

7. Nyakarahuka L, Tweheyo R. Occurrence and mortality rates from rabies in Uganda, 2001-2009. International Journal of Infectious Diseases 2012;16:e458.

8. Masiira B, Makumbi I, Matovu JKB, Ario AR, Nabukenya I, Kihembo C, et al. Long term trends and spatial distribution of animal bite injuries and deaths due to human rabies infection in Uganda, 2001-2015. PloS One 2018;13(8):e0198568.

9. Fèvre EM, Kaboyo RW, Persson V, Edelsten M, Coleman PG, Cleaveland S. The epidemiology of animal bite injuries in Uganda and projections of the burden of rabies. Tropical medicine & international health 2005;10(8):790–8.

10. Twabela AT, Mweene AS, Masumu JM, Muma JB, Lombe BP, HC. Overview of Animal Rabies in Kinshasa Province in the Democratic Republic of Congo. PloS One 2016;11(4):e0150403.

11. Cleaveland S, Fevre EM, Kaare M, Coleman PG. Estimating human rabies mortality in the United Republic of Tanzania from dog bite injuries. Bulletin of the World Health Organization 2002;80:304–10.

12. Sambo M, Cleaveland S, Ferguson H, Lembo T, Simon C, Urassa H, et al. The burden of rabies in Tanzania and its impact on local communities. PLoS neglected tropical diseases 2013;7(11):e2510.

13. Depani SJ, Kennedy N, Mallewa M, Molyneux EM. Evidence of rise in rabies cases in Southern Malawi–better preventative measures are urgently required. Malawi Medical Journal 2012;24(3):61–4.

14. Weyer J, Szmyd-Potapczuk AV, Blumberg LH, Leman PA, Markotter W, Swanepoel R, et al. Epidemiology of human rabies in South Africa, 1983–2007. Virus research 2011;155(1):283–90.

15. Nel LH, Rupprecht CE. Emergence of lyssaviruses in the Old World: the case of Africa. In: Wildlife and Emerging Zoonotic Diseases: The Biology, Circumstances and Consequences of Cross-Species Transmission. Springer, 2007:161–93.

16. Nel LH, Markotter W. Lyssaviruses. Critical reviews in microbiology 2007;33(4):301–24.

17. Knobel D, Bessell P, Grover M, Reininghaus B. Rabies and Wildlife Conservation in Africa World Small Animal Veterinary Association World Congress Proceedings, 2014. https://www.vin.com/apputil/content/defaultadv1.aspx?id=7054642&pid=12886&. 2014. Accessed 10 June 2020

18. Sabeta CT, Rensburg DD, Phahladira B, Mohale D, Harrison-White RF, Esterhuyzen C, et al. Rabies of canid biotype in wild dog (Lycaon pictus) and spotted hyaena (Crocuta crocuta) in Madikwe Game Reserve, South Africa in 2014-2015: Diagnosis, possible origins and implications for control. Journal of the South African Veterinary Association 2018;89(1):1–13.

19. Banyard AC, McElhinney LM, Johnson N, Fooks AR. Introduction History of rabies control by vaccination. Revue scientifique et technique (International Office of Epizootics) 2018;37(2):305–22.

20. Banyard AC, Horton DL, Freuling C, Müller T, Fooks AR. Control and prevention of canine rabies: the need for building laboratory-based surveillance capacity. Antiviral Research 2013;98(3):357–64.

21. Horton DL, Ismail MZ, Siryan ES, Wali ARA, Ab-dulla HE, Wise E, et al. Rabies in Iraq: trends in human cases 2001–2010 and characterisation of animal rabies strains from Baghdad. PLoS neglected tropical diseases 2013;7(2):e2075.

22. Lembo T, Hampson K, Kaare MT, Ernest E, Knobel D, Kazwala RR, et al. The feasibility of canine rabies elimination in Africa: dispelling doubts with data. PLoS neglected tropical diseases 2010;4(2):e626.

23. Peters MDJ, Godfrey CM, Khalil H, McInerney P, Parker D, Soares CB. Guidance for conducting systematic scoping reviews. International journal of evidence-based healthcare 2015;13(3):141–6.

24. Moher D, Liberati A, Tetzlaff J, Altman DG, Prisma Group. Preferred reporting items for systematic reviews and meta-analyses: the PRISMA statement. PLoS med 2009;6(7):e1000097.

25. Aromataris E, Riitano D. Constructing a search strategy and searching for evidence. Am J Nurs 2014; 114(5):49–56.

